# Effect of socioeconomic and ethnic characteristics on COVID-19 infection: The case of the Ultra-Orthodox and the Arab communities in Israel

**DOI:** 10.1101/2020.05.25.20111575

**Authors:** M Saban, T Shachar, O Miron, R Wilf-Miron

## Abstract

**Background:** During infectious disease outbreaks the weakest communities are more vulnerable to the infection and its deleterious effects. In Israel, the Arab and Ultra-Orthodox Jewish communities have unique demographic and cultural characteristics that place them at risk for infection.

**Objective:** To examine the socioeconomic and ethnic differences in relation to COVID-19 testing, cases and deaths, and to analyze infection spread patterns in ethnically diverse communities.

**Methods:** Consecutive data on COVID-19 diagnostic testing, confirmed cases and deaths collected from March 31^st^ through May 1^st^, 2020 in 174 localities across Israel (84% of the population) were analyzed by socioeconomic ranking and ethnicity.

**Findings:** Tests were performed on 331,594 individuals (4·29% of the total population). Of those, 14,865 individuals (4·48%) were positive and 203 died (1·37% of confirmed cases).

The percentage of the population tested was 26% and the risk of testing positive was 2·16 times higher in the lowest, compared with the highest socioeconomic category. The proportion of confirmed cases was 4·96 times higher in the Jewish compared with the Arab population.

The rate of confirmed cases in 2 Ultra-Orthodox localities increased relatively early and quickly. Other Jewish and Arab localities showed consistently low rates of confirmed COVID-19 cases, regardless of socioeconomic ranking.

**Interpretation:** Culturally different communities reacted differently to the COVID-19 outbreak and to government measures, resulting in different outcomes. Therefore, socioeconomic and ethnic variables cannot fully explain communities’ reaction to the pandemic. Our findings stress the need for designing a culturally adapted approach for dealing with health crises.

## Introduction

Cultural, behavioral, and societal differences, including socioeconomic (SE) status, health seeking behaviors, and intergenerational cohabitation may affect COVID-19 spread.^1^ More deprived and less affluent communities are more vulnerable to the disease and its deleterious effects due to a higher prevalence of comorbidities such as obesity, diabetes and hypertension.^2^ Such populations are also characterized by low health literacy and less access to information channels, causing them to miss or misinterpret health authorities’ instructions and warnings.^3^

Ethnic and racial disparities related to COVID-19 infection and mortality have been reported: British intensive care data showed that rates of confirmed cases among Asian and black patients were almost double that of the proportion of these ethnic minorities in the UK.^4^ In the United States, African Americans are over-represented both in the number of cases and in the number of deaths. For example, 52% of COVID-19 cases and 69% of COVID-19 deaths in Chicago involve black individuals, although blacks account for 30% of the population.^5^

In addition, differences in socioeconomic status also affect COVID-19 mortality. In England and Wales mortality rates have been twice as high in deprived areas as compared with the least deprived.^6^ Israel is a multiethnic multicultural country characterized by wide social disparities and a high proportion of people living in poverty.^7^ Poverty is disproportionally prevalent among two population groups: the Arabs and the Ultra-Orthodox (Haredi) Jews.

The Israeli Arab population accounts for 21% of the total of 9.1 million inhabitants. This population is generally younger, with 43% of the population younger than 18 (median age, 22 years), compared with 28% in the Jewish population (median age, 34).^8^ In addition, compared with the Jewish population, household net income is lower among the Israeli Arab population, fewer people hold an academic degree and participate in the labor force. The average number of household family members is 4·9, compared with 3·4 in Jewish families. Crowded living (more than 2 members per room) characterizes 26·5% of the families, compared with 4·6% of Jewish families.^9^ There is a higher prevalence of smoking–related chronic lower respiratory disease among Arab men, while both genders have higher prevalence of diabetes, hypertension and cardiovascular disease, compared with the Israeli population. ^9^

The Ultra-Orthodox (Haredi) are Jews that live by Jewish law and tradition.^10^ Accounting for 12% of the Israeli population, this group is younger, has larger families and is characterized by residential crowding – 26% have more than 2 people per room, compared with 2% of other non-orthodox Israeli Jews.^11, 12^ They reside mainly in Ultra-Orthodox-only cities or in separate neighborhoods in mixed cities. Employment rates in this population are low and household incomes are the lowest among all Israeli population groups.

Despite the known direct correlation between socioeconomic (SE) status and health, most of the Ultra-Orthodox population report very good health status and have a relatively high life expectancy,^12^ attributed to their high social capital.

The first patient with a confirmed COVID-19 infection arrived in Israel from the ‘Diamond Princess’ on February 21^st^, 2020. On February 27^th^, the first community-acquired case was diagnosed, and on March 20^th^, the first COVID-19-realted death was documented. Until May 1^st^, 16,101 cases and 225 deaths have been documented nationally.

Following the call to identify and address the toll of the pandemic on diverse populations, including racial and ethnic minorities and the poor,^13, 14^ we aimed to examine the socioeconomic and ethnic differences in relation to COVID-19 testing, confirmed cases and deaths, and to analyze infection spread patterns in diverse communities across Israel.

## Methods

### Data Sources

Data were obtained from the Israeli Ministry of Health’s (MOH) open COVID-19 database (https://data.gov.il/dataset/covid-19), which includes information on 174 medium or large urban communities/localities (5000 inhabitants or more) and is updated daily.

The database contains the number of COVID-19 diagnostic tests performed, the number of confirmed cases (i.e., those that tested positive by real-time quantitative reverse-transcriptase polymerase-chain-reaction [qRT-PCR] assay), and deaths in Israel.

We linked each locality in the MOH database to its socioeconomic (SE) cluster. SE clusters are homogenous units on a scale of 1 (lowest) to 10 (highest) that are determined by the Central Bureau of Statistics (CBS) according to population demography, education, employment and standard of living.^15^ SE clusters are further grouped by the CBS into four categories based on common characteristics with 1 as the lowest SE ranking (clusters 1-3); 2 (clusters 4-5), 3 (clusters 6-7) and 4 - the highest SE ranking (clusters 8-10). Categories 1, 2, 3 and 4 account for 28·5%, 18·3%, 27·2% and 25·9%, respectively, of the Israeli population.^16^

### Data analysis

Our analysis included MOH data from March 31^st^ to May 1^st^, 2020 (31 consecutive days.

MOH data on tests performed, confirmed COVID-19 cases and case fatality rate (CFR, defined as the ratio between confirmed deaths and confirmed cases) were analyzed by SE categories, ethnicity (Arab vs Jewish) and religiosity (Ultra-Orthodox Jewish vs. non-Ultra-Orthodox Jewish). As after April 26^th^, 2020 mortality data and ethnicity were no longer reported at the localities level in the MOH database, after this date, deaths reported in each hospital were allocated to the SE category of the hospital’s catchment area.

For the comparison between the Jewish and Arab populations, only localities in which more than 90% of the population is Jewish or Arab were included. Mixed cities were excluded because there is no accurate method to identify ethnicity in these cities.

Descriptive and frequency statistical analyses were performed using Matlab version 2020a and Python 3.6.5 software.

**Role of the funding source:** The funder of the study had no role in study design, data collection, data analysis, data interpretation, or writing of the report. The corresponding author had full access to all the data in the study and had final responsibility for the decision to submit for publication.

## Results

The 174 communities included in the analysis, comprise a population of 7·72 million inhabitants, or 84·0% of the population of Israel. The largest category (51 localities, accounting for 42·9% of the study population) belongs to the lower-mid SE category (SE category 2).

Table 1 presents data on COVID-19 testing rate, confirmed cases and mortality rate by SE category. As of May 1^st^, 2020, COVID-19 tests were performed on 331,594 individuals (4·3% of the total population). Of those, 14,865 individuals (4·48%) were positive for the infection and 203 deaths were reported (1·37% of confirmed cases).

**Table 1.** COVID-19 tests, confirmed cases and mortality, by SE category

|  | *Category 1* | *Category 2* | *Category 3* | *Category 4* | *Total* |
| --- | --- | --- | --- | --- | --- |
| ***Population*** | 1,667,076 | 3,312,699 | 1,502,970 | 1,237,666 | 7,720,411 |
| ***Tested*** | 84,282 | 139,197 | 58,712 | 49,403 | 331,594 |
| ***% of population*** | (5.05%) | (4.2%) | (3.91%) | (3.99%) | (4.29%) |
| **Confirmed** | 5,263 | 6,718 | 1,453 | 1,431 | 14,865 |
| *% of tested* | (6·24%) | (4·83%) | (2·47%) | (2·89%) | (4·48%) |
| **Deaths** | 12 | 108 | 50 | 33 | 203 |
| *% of confirmed* | (0·23%) | (1·61%) | (3·44%) | (2·31%) | (1·37%) |
| **Death per 100K**<br>**population** | **0.72** | **3.26** | **3.27** | **2.67** | **2.63** |

The analysis showed that the proportion of confirmed cases gradually decreased from category 1 through 3, with a slight increase in category 4. The lowest SE category (1) had the highest percentage of population tested for COVID-19 (5·05%) - 1·26 times higher, compared with the highest SE category (in which 3·99% of the population in that category were tested). The risk of testing positive for COVID-19 was 2·16 times higher in the lowest, compared with the highest SE category (6·24% vs. 2·89%).

COVID-19-related deaths documented during the analysis period (n=203), represent an overall mortality of 2·63 cases per 100K inhabitants. Mortality rates were unevenly distributed among the SE rankings, with SE category 3 demonstrating the highest number of deaths per 100K population and SE category 1 – the lowest (3·27 vs. 0·72). Category 2 and 3 showed similar rates – 3·26 and 3·27 deaths per 100K population, respectively.

CFR gradually increased from category 1 (0·23%) through 3 (3·44%), with a decrease in category 4 (2·31%, Table 1).

Arab localities comprise the majority (80%) of the lowest SE clusters (1-3) and 40% of cluster 4. Their proportion in cluster 5-6 is less than 10%, and they are not represented at all in clusters 6-10.^9^ Therefore, only homogenous localities from clusters 1-4 were included in the ethnicity analysis. Of the total Israeli Arab population of 1·92 million, 1·09 million (56·9%) were included in the analysis.

Overall, there were 538 and 78 confirmed cases per 100k population among Jews and Arabs, respectively. The proportion of confirmed cases among those tested was 4·96 times higher among Jews compared with Arabs. Figure 1 compares the Jewish and Arab population at the low and mid-low SE clusters (1-4 out of 10). In SE clusters 2 and 3, Jews performed 1·12 and 2·52 more testes, respectively, than Arabs, while in SE cluster 4 Arabs performed 1·38 more tests than Jews. In all SE clusters, the rate of confirmed cases was greater among the Jewish localities compared with the Arab ones.

**Figure 1.**
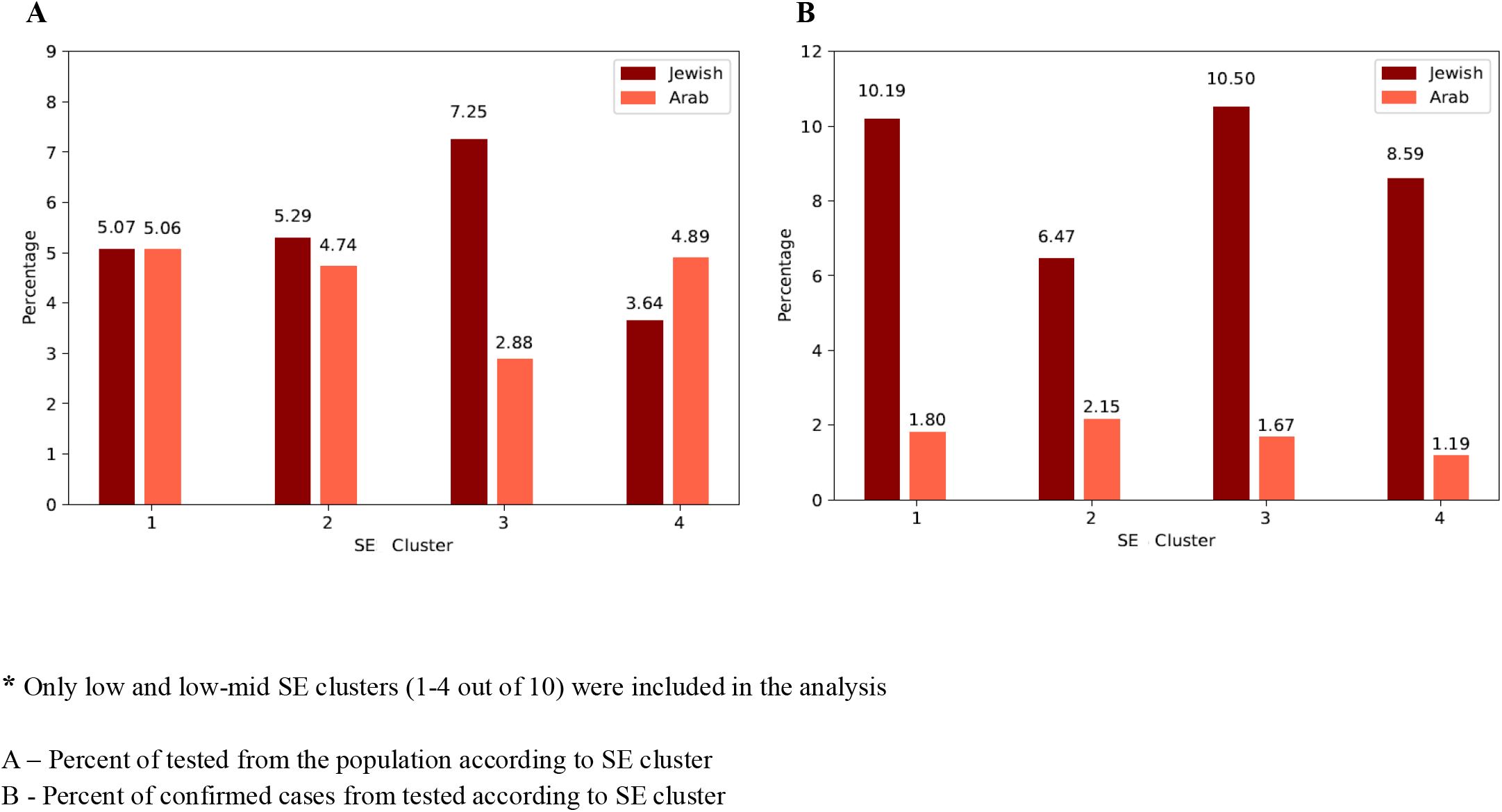
COVID-19 tests performed and confirmed cases by ethnicity and SE cluster. (A) Percent of tested from the population according to SE cluster. (B) Percent of confirmed cases from tested according to SE cluster. Only low and low-mid SE clusters (1-4 out of 10) were included in the analysis SES=socioeconomic status

To understand the epidemiology of COVID-19 by ethnicity and religiosity the proportion of confirmed COVID-19 cases was analyzed in six localities representing Arab, Ultra-Orthodox Jewish and non-Ultra-Orthodox populations each associated with a different SE category (Table 2). Bene-Beraq and El’ad are two predomenantly Ultra-Orthodox cities with 195,298 and 47,600 inhabitants, respectively, which belong to the lowest SE category (1). As shown in Figure 2, the rate of confirmed COVID-19 cases in these 2 cities increased relatively early and quickly (reflected in a steep slope). In comparison, Hertzliya, Beer Sheba (both predominantly Jewish non-Ultra-Orthodox cities) and the predominantly Arab city, Nazareth showed consistently low rates of confirmed COVID-19 cases during the entire period. Interestingly, Hertzliya, which belongs to the highest SE category, showed higher rates of cofirmed cases compared with Beer Sheba and Nazareth which are associated with SE categories 1 and 2, respectively. Hura, a Bedouin Arab locality, which is among the poorest communities in Israel (SE cluser 1 out of 10), demonstrated, a dramatic, rather late increase in confirmed cases, reaching the level of Bene Beraq, with its highest percentage of confirmed cases within a period of just 7 days.

**Table 2.** Proportion of COVID-19 tests and confirmed cases in selected Israeli localities from diverse SE categories*, cultural and religious backgrounds

|  | **Bene**<br>**Beraq** | **Beer**<br>**Sheba** | **Herzliya** | **Nazareth** | **El'ad** | **Hura** |
| --- | --- | --- | --- | --- | --- | --- |
| **SE Category** | 1 | 2 | 4 | 1 | 1 | 1 |
| Population | 195,298 | 196,755 | 90,900 | 78,252 | 47,600 | 16,983 |
| Tested<br>(% of population) | 24,685<br>(12.6%) | 8,277<br>(4.2%) | 3,508<br>(3.86%) | 1,288<br>(1.64%) | 4,069<br>(8.55%) | 769<br>(4.53%) |
| Confirmed<br>(% of tested) | 2,831<br>(11.4%) | 173<br>(2.09%) | 102<br>(2.9%) | 20<br>(1.55%) | 383<br>(9.41%) | 82<br>(10.66%) |
| Confirmed % of<br>population | 1.45% | 0.09% | 0.11% | 0.03% | 0.80% | 0.48% |

\*SE category: (1) = clusters 1,2,3; (2) = clusters 5-4; (4) = clusters 8,9,10

**Figure 2.**
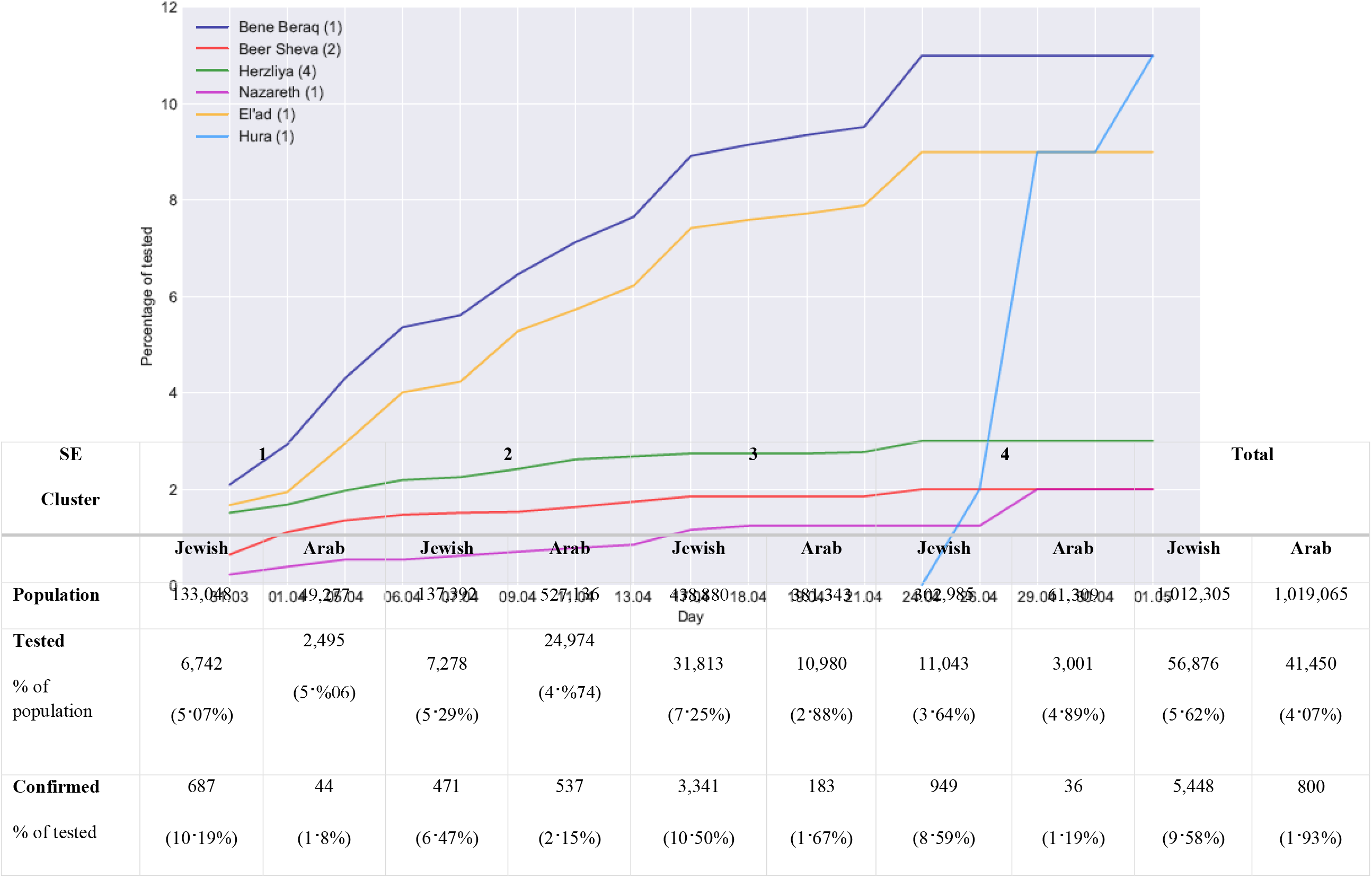
Proportion of confirmed COVID-19 cases in selected Israeli localities: Trends over a 31-day period. The percentage of accumulated confirmed COVID-19 cases out of those tested in selected localities between March 31^st^ and May 1^st^. The number in parentheses next to each locality’s name indicates SE category: (1) = clusters 1,2,3; (2) = 5-4; (3) = 6-7; (4) =8,9,10. Bene Beraq and El’ad are predominantly ultra-orthodox communities; Nazareth, and Hura are Arab cities while Herzliya and Beer Sheva are predominantly non-orthodox Jewish cities.

## Discussion

Our findings demonstrate the effects of socioeconomic, ethnic and cultural determinants on the epidemiology of COVID-19 in Israel.

Our analysis shows that more tests per population were performed in the most deprived populations compared with the least deprived ones. This might reflect the early awareness of the MOH to communities with high prevalence of confirmed cases, particularly the Ultra-Orthodox communities. As a result, epidemiological investigations and testing were directed to those communities.

A striking finding is the increasing rate of confirmed cases with decreasing SE categories, with rates more than double in the lowest category compared with the higher ones. A similar social gradient in health has been documented in New York City (NYC), where the likelihood that a test was positive was greater in poor neighborhoods, as well as in neighborhoods with larger households or a predominantly black population.^17^

CFRs were lower in low SE categories compared with higher SE categories. Due to the small number of COVID-19-related deaths in Israel, mortality data should be interpreted with caution. CFR in the Israeli population that was analyzed here (7·7 Million) was 1·37%, compared with 4·2%, 5·8% and 13·7% in Germany, US and Italy, respectively.^18^ Possible explanations for the relatively low mortality rate observed in Israel include the country’s good healthcare system with universal coverage of care,^19^ its younger population compared with other OECD members,^20^ which may be less affected by COVID-19,^21^ and early establishment of national measures. These measures included stopping of incoming flights and required 14-day isolation of those who arrived from abroad, social distancing measures, with an emphasis self-isolation of the elderly, a national state of emergency and almost complete lockdown between March 19^th^ and May 3^rd^, 2020. In addition, frequent military operations and emergency situations have prepared the Israeli healthcare system for crises, contributing to its ability to effectively care for severely ill patients.

As reported for other countries,^22^ it is possible that the number of COVID-19-related deaths in Israel have been under-estimated due to deaths that occurred at home prior to a COVID-19 diagnosis. During the first weeks of the pandemic, all health maintenance organizations in Israel advised their members to shift to telehealth platforms as the main venue for communicating with their health providers, instead of visiting their clinics. Since the most deprived patients are less comfortable with digital infrastructures,^23^ as well as less health literate, they might be more vulnerable to disease deterioration. However, as all Israeli citizens have access to public healthcare,^19^ the possibility of major under-reporting of infection and death is unlikely. Moreover, during the study period, all-cause mortality, reported by the Israel Center for Disease Control, was within range of the multiannual average,^24^ supporting this assumption.

Experience gained in prior health crises as well as from emerging reports from the COVID-19 pandemic, showed that ethnic minorities and depraved populations are more susceptible to being affected by the disease.^4, 5^ Therefore it was expected that the Arab ethnic minority in Israel, which is demographically characterized by being less affluent, having larger families, greater population density, residence in geographical peripheries, and a higher prevalence of co-morbidities, would demonstrate higher infection rates and more severe disease course compared with the general population. However, only moderate differences in testing were noted between the Jewish and Arab populations (5·6% vs. 4·1%), which might be explained by less access to healthcare services in a population of which more than half reside in geographical peripheries^9^ and fear of stigma and social, cultural and economic consequences of being found “positive”. To help overcome these obstacles, political representatives urged the MOH to increase testing among the Arab population.

The strikingly low infection rates among the Arab population – almost five times lower compared with the Jewish population – may be attributed to the following: a) Obedience of the population, which is predominantly (85%) Muslim^9^ and religious, to the official religious instructions (named “Fatwa” in Arabic) conveyed by the religious leaders (“Mufti”). The religious leaders instructed the population to follow the instructions of the MOH and relevant official bodies; to postpone or shorten cultural events; to perform funeral services with minimal participation and to avoid any kind of gathering together.^25^ They also closed all of the mosques, instructing people to pray at home instead, especially during Ramadan, which began on April 23rd. b) Arab men and women are employed at high rates in the Israeli healthcare system as physicians, nurses and pharmacists - the most prominent occupations of the COVID-19 crisis – contributing to the feeling that the Israeli Arabs citizens are full partners in this struggle.^9, 26^ This high rate of knowledgeable people could possibly contribute to maintaining social distancing and other measures to fight the disease. c) Traditionally, Arab families take care of the elderly at home and do not place them in residential care facilities, reflecting a culture that is more tolerant toward older adults.^27,28^ This may have been a protective factor, as a third of all COVID-19 deaths in Israel during the analysis period were documented among elderly living in residential homes.^29^ d) Finally, the political alliance of the main Arab-majority political parties in Israel – the Joint List – showed involvement by focusing on issues relating to all Israeli vulnerable sectors.^26^ Their voice was loudly heard among their voters and among policy makers.

Despite what seems to be an optimistic situation among the Israeli Arabs, one must consider the possibilities of clusters of undiagnosed infections, particularly in remote, segregated communities that are isolated geographically. Lower access to healthcare services might also explain late testing, and consequently a relatively late explosion of cases. The last week of April and the first days of May, saw a surge of confirmed cases in some Arab localities, such as the Bedouin locality of Hura (Figure 2); however, it is too early to judge if the Moslem Arabs followed their leaders’ instructions to avoid large gatherings during the month-long Ramadan holiday. Such gatherings may reverse the seemingly lower prevalence of COVID-19 demonstrated in the Arab society during the first month of widespread disease in Israel.

The presentation of COVID-19 in the Ultra-Orthodox community, which bears socioeconomic similarities to the Arab community, was completely different, manifesting early with very high infection rates, as shown in the two localities that are predominantly Ultra-Orthodox – Bene Beraq and El’ad (Figure 2). The community participates only minimally in secular affairs and opposes features of public Israeli life, such as the national government and modern law. Most Ultra-Orthodox people perceive an external threat from the surrounding secular society and are in constant need to react to and defend themselves from such threats. This translates into segregation from the rest of Israeli society.^30^ The emergency orders and restrictions issued by the government hit the most fundamental components of their identity – group studies of the bible (Torah), prayer in a quorum of ten men and ritual baths. These traditions were stronger than the biblical rule that everything should be put aside for the sake of saving lives.^26^ The instructions for strict social distancing were rejected because they were not endorsed by the community’s own leaders. Moreover - the senior leader of this community loudly opposed the instructions to close schools and told his followers to continue group learning. These Ultra-Orthodox leaders stopped opposing the government’s instructions only after massive infection spread was seen in their communities. To curb the spread of infection and flatten the escalating curve, the Israeli government imposed hermetic quarantine on some Ultra-Orthodox neighborhoods.

Our analysis has several limitations: First, data were analyzed by locality, rather than by individuals. This ecologic approach misses possible differences in the composition and disease patterns within localities. Second, small towns and settlements, comprising less than 5000 residents, were not included in the MOH database and therefore were not analyzed. No data exists to evaluate this possible bias on disease measures. Third, data on localities comprising mixed Arab-Jewish ethnicities were excluded from the analysis, because it was not possible to distinguish between the ethnicity of individuals in the database. Arab communities in large mixed cities might present with different disease patterns. Last, after April 26^th^, 2020 mortality data and ethnicity were no longer reported at the localities level in the MOH database; however, due to intensive media coverage of fatalities, including the naming of most of those who died, we believe that large numbers of Arab deaths could have not escaped public awareness.

Our analysis shows that we should treat assumptions about ethnic minorities with caution as it is not always possible to predict how such communities would react in times of crisis. The Israeli Arab community mostly demonstrated responsible behavior, following governmental instruction that were mediated by the community’s religious leaders. Although, the Ultra-Orthodox community did not abide by the government’s instructions because it regards its religious leaders’ instructions as more relevant, it is important not to stigmatize a community “en-bloc”. Such labeling of the whole community is unjust and counterproductive. Instead, the more cooperative leaders of the community should be strengthened so that their voice may be heard.

## Conclusions

Some lessons learnt from our analysis might be of value for the international audience. First, researchers and policymakers should adopt “tailor-made” perspectives and measures for dealing with culturally different communities during epidemics. Specifically, the cases presented here, which showed that the reactions to the crisis of two culturally different communities from similar socioeconomic backgrounds resulted in completely different outcomes, emphasize the need for designing a culturally adapted approach to curb viral spread. Second, our data prove that socioeconomic and ethnic variables cannot fully forecast and explain communities’ reaction to a crisis. The case of Nazareth, an Arab city with a deprived population that had lower infection rates compared with Herzliya, which is among the least deprived Jewish cities, is one example. Last, but not least, the collaboration of community leaders - whether religious or political representatives of the community – is essential for helping to change the course of the infection in their communities.

### Declaration of interests

The authors declare no conflict of interest. This study is solely submitted to publication in Lancet public health.

This research received no specific grant from any funding agency in the public, commercial, or not-for-profit sectors.

All authors have read and agreed to the content of the manuscript and confirmed the ICMJE.org authorship criteria.

## Data Availability

All data available

https://govextra.gov.il/ministry-of-health/corona/corona-virus/

## Acknowledgments

None.

## References

1. Pareek M, Bangash MN, Pareek N, et al. Ethnicity and COVID-19: an urgent public health research priority. Lancet 2020; 395(10234): 1421–2.

2. Allen L, Williams J, Townsend N, et al. Socioeconomic status and non-communicable disease behavioural risk factors in low-income and lower-middle-income countries: a systematic review. Lancet Glob Health 2017; 5(3): e277-e89.

3. Pirisi A. Low health literacy prevents equal access to care. Lancet 2000; 356(9244): 1828.

4. Rimmer A. Covid-19: Disproportionate impact on ethnic minority healthcare workers will be explored by government. BMJ 2020; 369: m1562.

5. Yancy CW. COVID-19 and African Americans. JAMA 2020.

6. Deaths involving COVID-19 by local area and socioeconomic deprivation: deaths occurring between 1 March and 17 April 2020: Office for National Statistics, 2020.

7. Bleikh H. Poverty and Inequality in Israel: Trends and Decompositions Jerusalem: Taub Center for Social Policy Studies in Israel, 2016.

8. Weiss A. The Singer Series: State of the Nation Report 2018. Jerusalem: Taub Center for Social Policy Studies in Israel, 2018.

9. Chernichovsky D, Bisharat B, Bowers L, Brill A, Sharony C. The Health of the Arab Israeli Population. Jerusalem: Taub Center for Social Policy Studies in Israel, 2017.

10. Schnall E. Multicultural Counseling and the Orthodox Jew. Journal of Counseling & Development 2006; 84(3): 276–82.

11. Cahaner L, Malach G. The Yearbook of the Ultra-Orthodox Society in Israel. Jerusalem: The Israel Democracy Institute, 2019.

12. Chernichovsky D, Sharony C. The Relationship Between Social Capital and Health in the Haredi Sector. Jerusalem: Taub Center for Social Policy Studies in Israel, 2015.

13. Haynes N, Cooper LA, Albert MA, Association of Black C. At the Heart of the Matter: Unmasking and Addressing COVID-19’s Toll on Diverse Populations. Circulation 2020.

14. Laurencin CT, McClinton A. The COVID-19 Pandemic: a Call to Action to Identify and Address Racial and Ethnic Disparities. J Racial Ethn Health Disparities 2020.

15. Characterization and Classification of Geographical Units by the Socio-Economic Level of the Population Jerusalem: Central Bureau of Statistics, 2015.

16. The national protam for quality indicators in community healtcare. Report for the yeasr 2016–2018.: The Ministry of Health and the Naitonal Institue for Health Policy Research, 2019.

17. Borjas GJ. Demographic Determinants of Testing Incidence and COVID-19 Infections in New York City Neighborhoods. National Bureau of Economic Research Working Paper Series 2020; 26952.

18. Johns Hopkins University and Medicine. Coronavirus resource center. World map. 2020. https://coronavirus.jhu.edu/map.html.

19. OECD. OECD Reviews of Health Care Quality: Israel 2012; 2012.

20. OECD. Elderly population. 2014.

21. Ruan Q, Yang K, Wang W, Jiang L, Song J. Clinical predictors of mortality due to COVID-19 based on an analysis of data of 150 patients from Wuhan, China. Intensive Care Med 2020.

22. Gaye B, Fanidi A, Jouven X. Denominator matters in estimating COVID-19 mortality rates. Eur Heart J 2020.

23. Armenta A, Serrano A, Cabrera M, Conte R. The new digital divide: the confluence of broadband penetration, sustainable development, technology adoption and community participation. Information Technology for Development 2012; 18(4): 345–53.

24. Monitoring of COVID-19 in Israel: Report for the week that ends in 2 may 2020.: Ministry of Health, 2020.

25. The house of Fatwa and Islamic Research in the Palsetinian Interior 48. 2020. https://www.facebook.com/DarEftaa48/.

26. Sheleg Y. The Coronavirus and Israel’s Ultra-Orthodox and Arab Communities. Jerusalem: The Israeli Democracy Institute, 2020.

27. Bergman YS, Bodner E, Cohen-Fridel S. Cross-cultural ageism: ageism and attitudes toward aging among Jews and Arabs in Israel. Int Psychogeriatr 2013; 25(1): 6–15.

28. Suleiman K, Walter-Ginzburg A. A nursing home in Arab-Israeli society: targeting utilization in a changing social and economic environment. J Am Geriatr Soc 2005; 53(1): 152–7.

29. Yasour Beit-Or M. Dr. Vered Ezra will replace Professor Gamzo in dealing with Nursing homes. Israel Ha’yom. 2020 4 May 2020.

30. Sivan E, Caplan K. Israeli Haredim: integration Without Assimilation?: The Van Leer Jerusalem Institute and Hakibbutz Hameuchad; 2003.

